# Colorectal cancer screening in the face of constrained resources and the emerging epidemic among the young

**DOI:** 10.1101/2025.07.24.25332117

**Authors:** Brian Cox, Phil Bagshaw, John D. Potter

## Abstract

We assessed screening strategies to ameliorate the increasing incidence of colorectal cancer (CRC) among those <50 years in Aotearoa New Zealand (ANZ). We analysed CRC incidence and mortality and estimated the cost-effectiveness of screening with: one-off flexible sigmoidoscopy (FS) at age 40 (FS40) and 45 (FS45); and faecal immunochemical testing (FIT) for with a sensitivity cut-off of 75ng Hb/ml buffer (FIT75) and FIT200.

CRC incidence and mortality varied across birth cohorts, reached a low point for those born in the 1960s, and rose again thereafter. FS40 and FS45 would prevent >50% of CRC deaths over 20 years, vs. approximately 13% for FIT75 or FIT200. FS45 was shown to be the most effective and cost-effective strategy.

Long-term benefit would accrue from the adoption of FS screening and, where needed, FS diagnosis. Establishment of an FS-training course for nurses and other healthcare personnel would expand the required service provision.

## Introduction

The lifetime risk of colorectal cancer (CRC) for any specific age in Aotearoa New Zealand (ANZ),^1^ as in some other cuntries,^2^ is influenced by the generation into which a person was born (birth cohort). Among those <50 years of age, born in the mid 1970s onward, the incidence of CRC has increased.^2,3^ There are important differences between the presentation of this young-onset CRC (YOCRC) and the presentation of CRC in older people. About 80% of YOCRC occurs in the descending colon and rectum (L-sided), 70% present with stage 3 or 4 disease, have better overall survival than expected, but lower survival if they have R-sided CRC. The majority of YOCRC are of unknown aetiology, although there is some evidence of an association with known risk factors.^4^

Detection using a biennial faecal immunochemical test (FIT) is influenced by the cut-off value chosen.^5^ The choice of the value (usually set from 10ng to 200ng Hb/ml of buffer) identifies a small high-risk subpopulation for whom the likely benefit of screening by colonoscopy exceeds the risks of the procedure.

Direct inspection of the descending colon and rectum by flexible sigmoidoscopy (FS), usually reaches the splenic flexure. FS requires considerably less time to complete than colonoscopy and is the most efficient initial endoscopic examination for people presenting with “outlet” rectal bleeding or persistent anaemia.^6–8^ Some of the abnormalities found during L-sided endoscopy are good indicators of whether subsequent completion colonoscopy is indicated,^6,7,9^ and are assisted by testing for iron-deficiency anaemia.^10^

Screening of the asymptomatic population from 45 years of age has been suggested, in ANZ and elsewhere, as a response to the increase in YOCRC mortality.^11^ The primary aim of screening for disease is to reduce mortality.^12^ Screening by colonoscopy has been shown to be the most effective method of reducing CRC incidence and mortality^13^ but major complications can occur, which hinder its use as a screening test. Less burdensome one-off FS has repeatedly been identified as a cost-effective approach for CRC screening.^14,15^

The effects and costs of screening and treatment from reducing the starting age of the National Bowel Screening Programme (NBSP); reducing the cut-off value from 200 to 75ng/ml; or introducing one-off FS at 40 or 45 years of age were estimated. The projected CRC mortality and incidence and the costs of screening scenarios were estimated, as were the savings made: by avoiding the costs of treatment; in the taxation not lost because of premature death; and in not needing to pay survivor benefits.

## Methods

The <u>annual number of deaths</u> in ANZ by sex and age for subsites of CRC from 2000 to 2022, CRC incidence data from 2000 to 2017 and estimates of the <u>past and projected</u> <u>population</u> were used to predict future age-specific CRC mortality and incidence. The introduction of the NBSP for people 60-74 years of age may have altered the presentation and diagnostic pathway for YOCRC; therefore, only CRC incidence up to 2017, the beginning of the NBSP, was analysed.

In 2014, there was a sudden increase in registrations of neuroendocrine carcinoid tumours of the appendix by the New Zealand Cancer Registry which were not previous classified as invasive. To avoid misclassification bias in the analysis, registrations with the ICD-10 rubric C181 (cancer of the appendix) were excluded.

Trends in incidence and mortality rates were assessed by age-cohort Poisson models using Stata^TM^ adjusted for sex and the ratio of Māori to non-Māori ethnicity. Changes in risk of CRC for each birth cohort: (i) relative to the risk of those born about 1958 for incidence, and (ii) risk of death from CRC relative to those born about 1957 were examined. Projections of CRC mortality and incidence rates included birth cohort differences in risk.^17^

The potential effect of extending screening to start at 40 or 45 years of age in 2027 was assessed. The rate of detection of R-sided abnormalities was set at 40% below the detection rate of L-sided disease for both the FIT75 and FIT200 scenarios. Per-protocol estimates of the effects of FIT and FS on CRC incidence and mortality from randomised controlled trials were applied to the proportion of the population screened. The projected annual population of cohorts of men and women from age 40 and 45 in 2027 provided estimates of the annual population who may benefit from screening starting in 2027. Five-year incidence and mortality rates were used to estimate the expected number of CRC cases and deaths for those 40 and 45 years of age in 2027. The effects of the sex ratio, site distribution, and extent of disease of those who developed YOCRC, born 1978-1986, was also assessed.

### The impact on CRC of screening with one-off FS or 2-yearly FIT

The ratio of R-sided to L-sided YOCRC is lower than for CRC at older ages, which required the effects of each screening scenario to be calculated separately for L-sided and R-sided tumours. For one-off FS screening, 5% were expected to be referred for completion colonoscopy^17^ and 95% of those referred were assumed to undergo the procedure. Per-protocol estimates of the effect of one-off FS on the incidence of R-sided CRC for each year of follow-up were used (Figure 4 of supplementary material in Wooldrage et al 2024^17^) to calculate the expected annual number requiring treatment following screening participation starting at 40 years (FS40) or 45 years of age (FS45). The proportional reduction in L-sided CRC mortality from one-off FS for those aged 55-59 years of age was estimated to be, 75% for men and 60% for women.^17^

Annual CRC mortality and incidence prevented by FIT75 and FIT200 screening were estimated. Comparisons of the sensitivity for detection of CRC and advanced adenoma were restricted to the test performance at different cut-off levels of the OC-Sensor FIT wherever possible.

The annual hazard rate ratios (HRR) for CRC incidence and mortality from biennial screening with gFOBT in the Nottingham screening trial^18^ were adjusted for a FIT75 to gFOBT detection ratio (DR) of 1.5.^19^ Therefore, the per-protocol HRR of FIT75=1+((per protocol HRR of gFOBT-1)×DR). The participation in the screening arm of the Nottingham trial was 78% and the estimated FIT per-protocol HRR was adjusted by (per-protocol HRR-0.22)÷0.78.

The HRR of FIT75 was adjusted in a similar manner using the detection ratio of FIT200 to FIT75 for CRC of 0.79;^5^ i.e., 2-yearly FIT200 was estimated to detect 21% fewer cases of CRC than 2-yearly FIT75, similar to the 18% difference predicted by the ANZ pilot study.^20^ The same adjustment was made for the predicted prevention of mortality by FIT200 compared to FIT75. For all screening modalities, it was assumed that 95% of those referred for colonoscopy would accept it and, of those, 25% would require 3-yearly surveillance by colonoscopy.

### Health costs of screening scenarios to reduce the impact of rising rates of CRC

For each screening scenario, patients who underwent colonoscopy were removed from participation in subsequent rounds of screening. The cost of hemicolectomy for CRC was estimated to be $17,090,^21^ chemotherapy NZ$25,000, and average cost of treatment NZ$42,095. The cost of the <u>investigation of symptoms by colonoscopy</u> was set at NZ$2,895 and by FS NZ$1,794. The cost of collection and reporting of pathology specimens was included. The cost of a <u>faecal immunochemical test (FIT) to investigate</u> <u>symptoms</u> was set at NZ$132. An annual discount rate of 2% was applied to costs and events in the year they occurred.

The discounted health-service savings from treatment avoided because of reduced incidence were subtracted from the cost of screening and subsequent colonoscopies and divided by the discounted number of deaths from CRC prevented to estimate the ratio of the health-service costs of screening and treatment per death prevented.

### Other government costs

The government taxation foregone from early death from CRC in people 40-54 years of age was estimated from <u>average income and proportion of men and women in</u> <u>employment in 2024</u>. Individual income in men and women is <u>highest in this age group</u> with men contributing about twice as much as women per capita.^22^ Including those without direct income, the average direct tax per person 40-64 years of age in 2023 was NZ$5,815. The estimate of total tax assumed that 70% of earnings after direct tax was used for the purchase of goods and services incurring 15% GST. The total average government tax was estimated to be NZ$11,530 per person, 40-64 years of age.

Eighty per cent of deaths from CRC were assumed to result in the payment of a survivor’s benefit for 2 years, The annual cost of the survivor’s benefit in 2024 was: NZ$31,837; NZ$34,571; NZ$34,710; NZ$32,786; and NZ$29,126, respectively, for the 40-44, 45-49, 50-54, 55-59, and 60-64 year age groups. This was discounted by 2% per annum.

## Results

### Generation risks of CRC

The risk of CRC mortality and incidence for different generations varied (p<0.001) and the risk of death relative to those born about 1957, and the risk of developing CRC relative to those born about 1958, are shown in Figure 1. Risk of CRC mortality for generations born about 1982 was 87% higher (95%CI:41%-147%) than those born about 1957.

**Figure 1.**
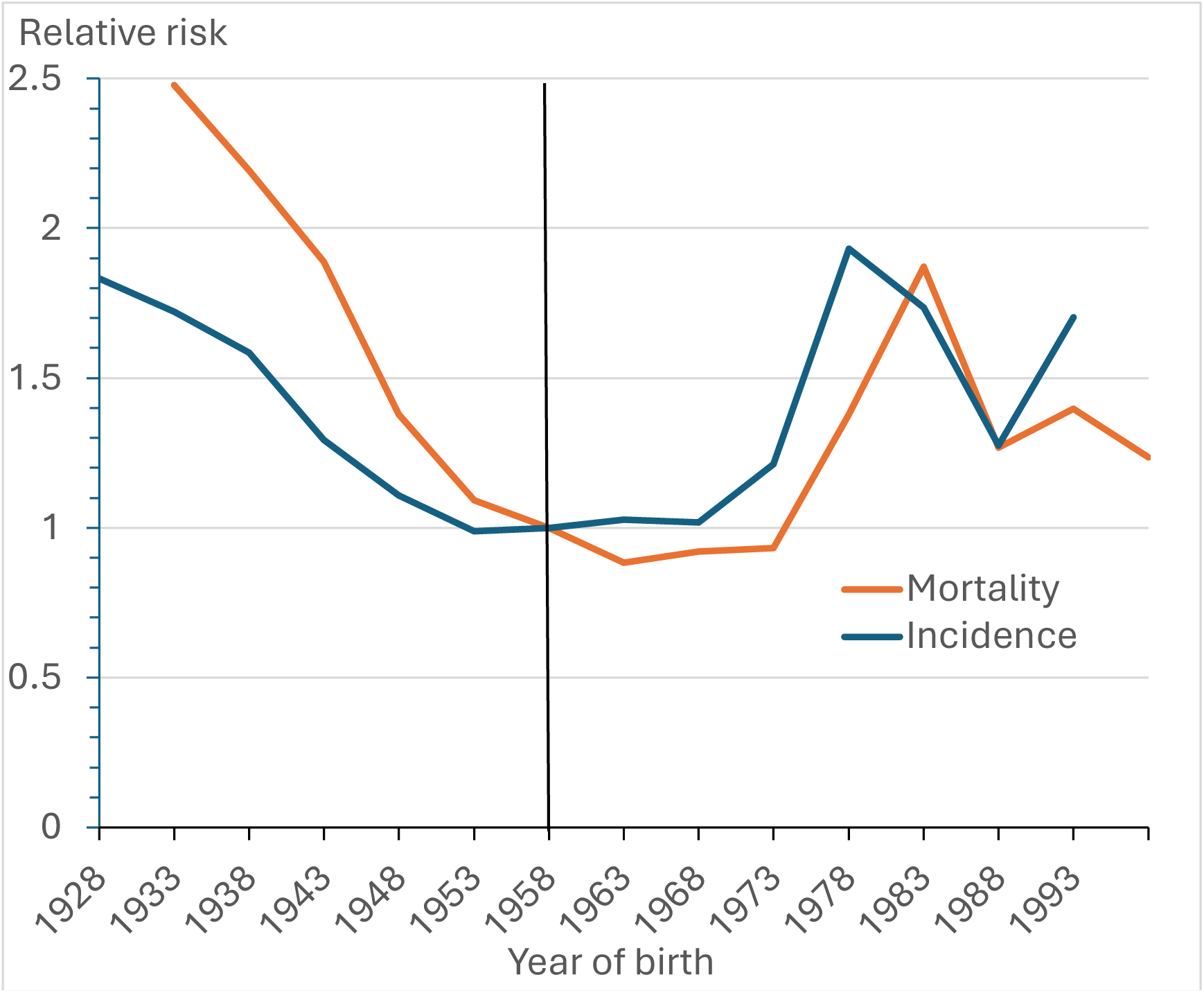
Risk of mortality and incidence from CRC for different generations compared to those born about 1957 and about 1958, respectively.

The reduced risk of developing CRC for those born in the mid-1950s^1^ persisted only up to generations born about 1968 before rising to become 93% higher (95%CI:58%-136%) for those born about 1978 relative to generations born 20 years earlier.

### Trends and prediction of CRC mortality 30-59 years of age

The CRC mortality in ANZ for different 5-year age groups in those aged <60 years for the 2002-06, 2007-11, 2012-16, and 2017-21 time-period and projected mortality is shown in Table 1. The sum of mortality rates for those aged 30-34 years in 2007-11, 35-39 years in 2012-16, and 40-44 years in 2017-21, multiplied by the length of the age groups, estimates the probability of people born about 1977 dying from CRC aged 35-49 years; i.e., their risk was (1.3+3.6+7.1)×5=60.0 per 100,000. The comparable risk for those born 5 years earlier was (1.0+2.7+4.9)×5=43.0 per 100,000. Thus, the risk of CRC mortality aged 30-44 years was 40% higher for people born around 1977 than for those born around 1972. Mortality rates aged 35-44 years for those born about 1982 were 84% higher than for those born 10 years earlier.

**Table 1.**
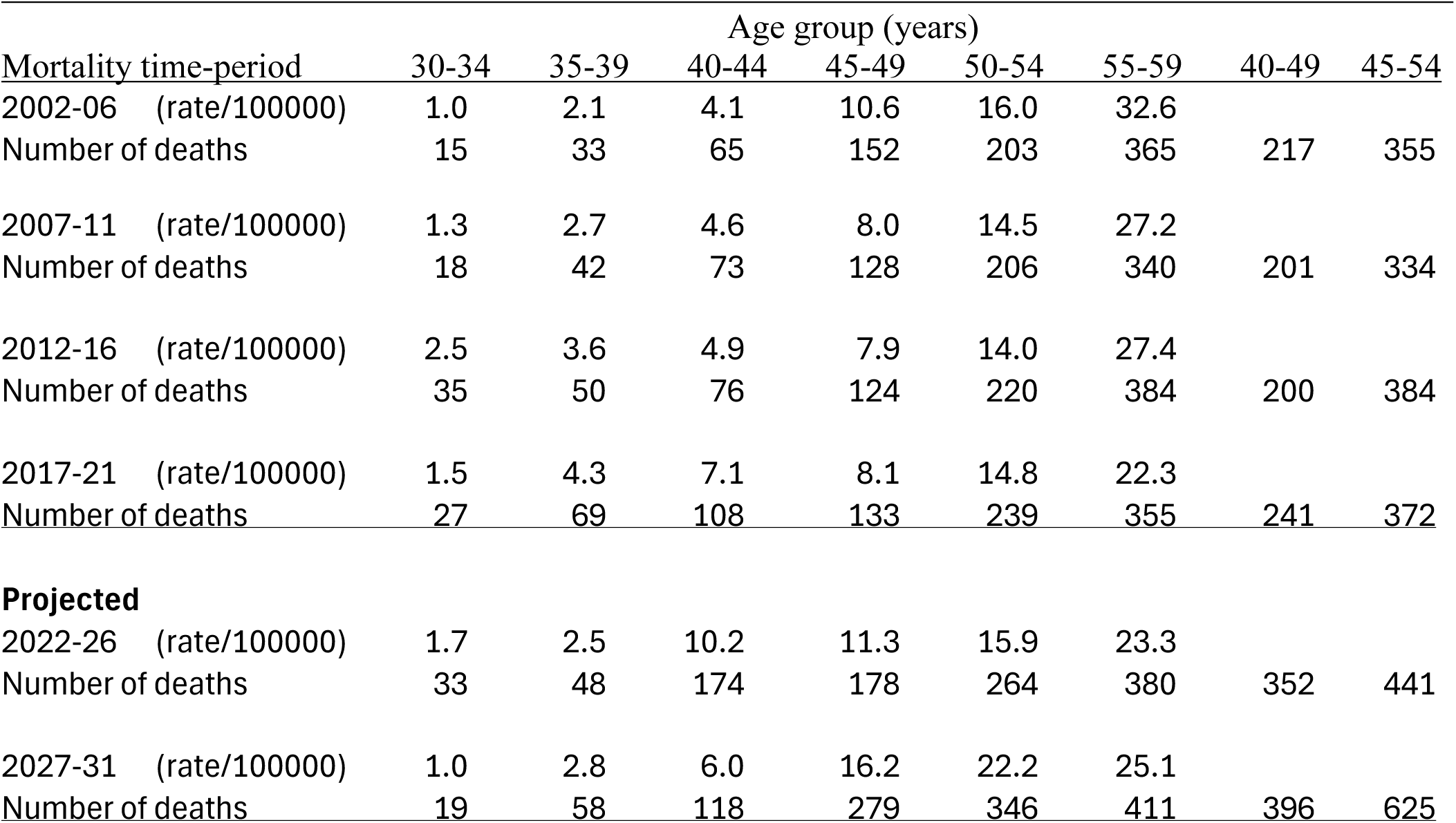
Overall CRC mortality for younger age groups and projected mortality to the 2027-31 time-period.

**Table 2.**
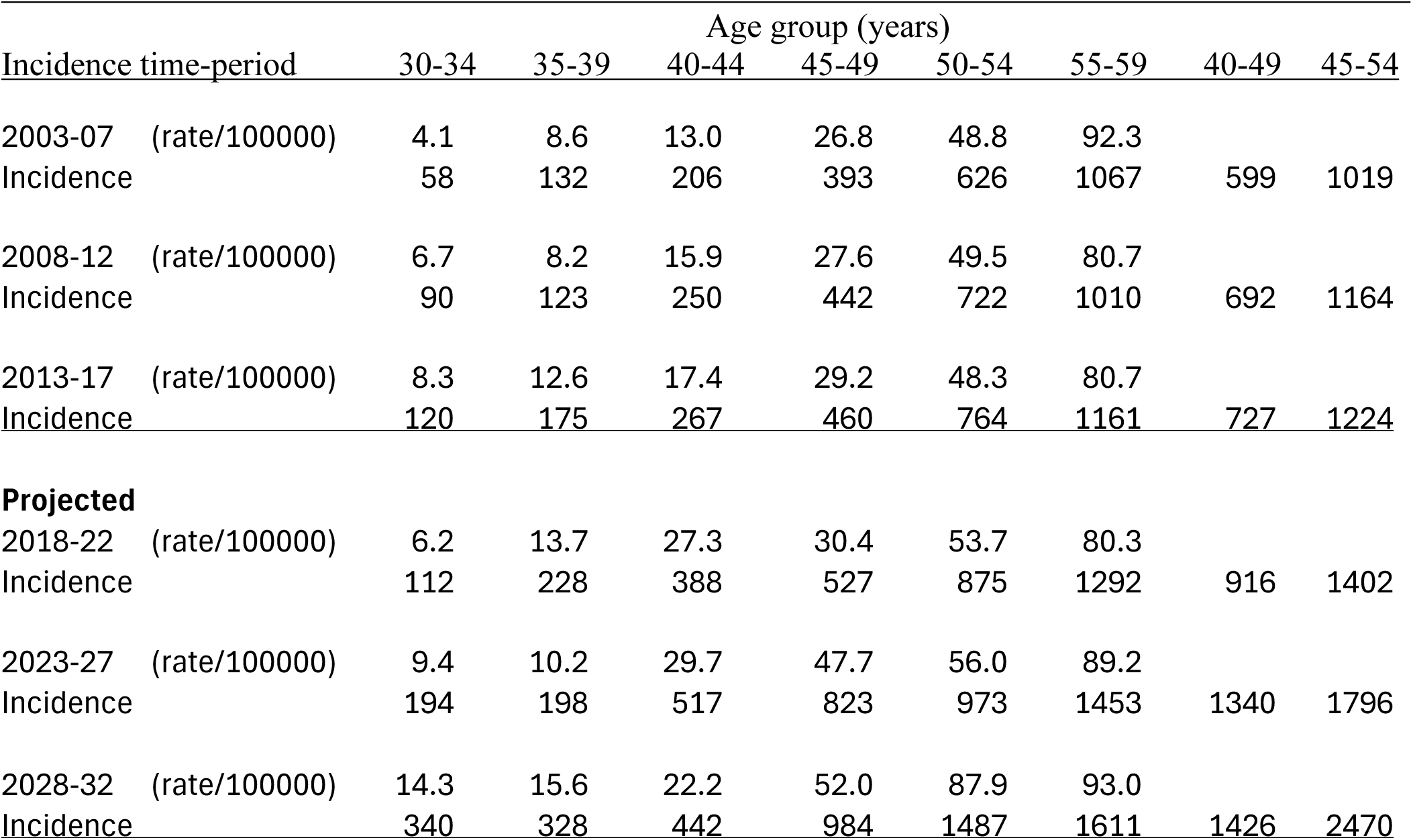
Overall CRC incidence, excluding cancer of the appendix, for younger age groups and projected incidence to the 2028-32 time-period.

CRC mortality in people under 50 years of age is expected to increase in the 2027-31 time-period to 79 deaths per year for those 40-49 years of age and 105 deaths per year for those 45-54 years of age. Poisson regression of CRC mortality established significant differences in risk for different birth cohorts (P<0.0001). The lifetime rate ratio (RR) of mortality of those born about 1982 compared to 1957, adjusted for sex and ethnicity, was 1.86 (95%CI:1.41-1.25). This effect was most pronounced for non-Māori men (RR=2.54;95%CI:1.68-3.83).

### Trends and prediction of CRC incidence 30-59 years of age

Incidence rates for those aged 30-34 years in 2008-12 and 35-39 years in 2013-17, born about 1978, were on average 57% higher than those born 5-years earlier. The risk of developing CRC for differed for birth cohorts (p<0.0001). The lifetime risk for those born about 1978 was 93% (95%CI:58-136%) higher than those born about 1958 (Figure 1). With adjustment for birth cohort, the male to female RR was 1.04 (95%CI:0.96-1.12) and the Māori to non-Māori RR was 0.99 (95%CI:0.88-1.10).

In 2013-17, the proportion of CRC in the 40-49 year age group reachable by FS was 80% for men and 76% for women, of which 69% and 80%, respectively, were not localised at diagnosis.

### FS40 screening

The estimated effectiveness of screening over 20 years using FS 40 years in 2027 (born in 1987) is shown in Table 3. Calculations were conducted separately for men and women but shown for the sexes combined. The projected male and female individual age-specific mortality and incidence rates were used to estimate the mortality and incidence expected without screening. Men have been shown to participate in FS screening slightly more frequently than women; hence, participation was chosen to be 55% for men and 50% for women. Over 20 years of screening, 138 (54%) of the expected 253 deaths were estimated to be preventable from one-off FS. Each prevented death required 356 members of the cohort to be screened.

**Table 3.** Twenty-year effect of screening by once-only flexible sigmoidoscopy at 40 years of age: participation 52.5% of which 5% recommended for colonoscopy of whom 25% require 3-yearly surveillance (costs given in NZ dollars).

| Age | Annual popn | Mort rate | Exp dths | Num FS | Colono-<br>scopies | Mort HR | Dths prev | Inc rate CRC | CRC w/o FS | Inc Distal HR | Cases treated | Disc cost of screening | Disc savings of treatment | Disc dths prev |
| --- | --- | --- | --- | --- | --- | --- | --- | --- | --- | --- | --- | --- | --- | --- |
| 40 | 71940 | 4.6 | 3 | 37775 | 1794 | 1 | 0 | 22.2 | 16 | 1.50 | 3 | \$72,962,885 | \$129,640 | 0.0 |
| 41 | 71870 | 5.4 | 4 | | | 0.87 | 1 | 26.1 | 19 | 0.21 | -6 | | -\$250,760 | 1.0 |
| 42 | 71810 | 6.2 | 4 | | | 0.74 | 2 | 30.0 | 22 | 0.12 | -8 | | -\$326,590 | 2.3 |
| 43 | 71740 | 7.0 | 5 | | 426 | 0.74 | 3 | 33.9 | 24 | 0.18 | -8 | \$1,233,702 | -\$325,721 | 2.5 |
| 44 | 71660 | 7.8 | 6 | | | 0.74 | 3 | 37.7 | 27 | 0.25 | -8 | | -\$317,537 | 2.8 |
| 45 | 71580 | 8.6 | 6 | | | 0.74 | 3 | 41.6 | 30 | 0.26 | -8 | | -\$316,134 | 3.0 |
| 46 | 71490 | 9.4 | 7 | | 405 | 0.74 | 4 | 45.5 | 33 | 0.20 | -10 | \$1,172,017 | -\$366,642 | 3.3 |
| 47 | 71390 | 10.2 | 7 | | | 0.74 | 4 | 49.4 | 35 | 0.20 | -11 | | -\$395,077 | 3.5 |
| 48 | 71300 | 12.5 | 9 | | | 0.74 | 5 | 60.1 | 43 | 0.24 | -13 | | -\$463,241 | 4.2 |
| 49 | 71200 | 14.9 | 11 | | 385 | 0.74 | 6 | 65.7 | 47 | 0.25 | -14 | \$1,113,416 | -\$488,751 | 4.9 |
| 50 | 71080 | 17.2 | 12 | | | 0.74 | 7 | 73.9 | 53 | 0.26 | -15 | | -\$526,861 | 5.5 |
| 51 | 70960 | 19.5 | 14 | | | 0.74 | 8 | 82.0 | 58 | 0.27 | -17 | | -\$566,123 | 6.2 |
| 52 | 70830 | 21.9 | 15 | | 365 | 0.74 | 9 | 90.2 | 64 | 0.28 | -17 | \$1,057,745 | -\$546,634 | 6.8 |
| 53 | 70700 | 24.1 | 17 | | | 0.74 | 10 | 107.1 | 76 | 0.31 | -21 | | -\$684,637 | 7.3 |
| 54 | 70560 | 26.4 | 19 | | | 0.74 | 10 | 124.0 | 88 | 0.29 | -23 | | -\$736,234 | 7.8 |
| 55 | 70410 | 28.7 | 20 | | 349 | 0.74 | 11 | 140.9 | 99 | 0.35 | -25 | \$1,004,858 | -\$779,404 | 8.3 |
| 56 | 70250 | 30.9 | 22 | | | 0.74 | 12 | 157.8 | 111 | 0.40 | -26 | | -\$802,549 | 8.8 |
| 57 | 70080 | 33.2 | 23 | | | 0.74 | 13 | 174.7 | 122 | 0.42 | -31 | | -\$936,907 | 9.2 |
| 58 | 69900 | 34.3 | 24 | | 332 | 0.74 | 13 | 192.3 | 134 | 0.38 | -35 | \$954,615 | -\$1,009,989 | 9.3 |
| 59 | 69720 | 35.4 | 25 | | | 0.74 | 14 | 209.8 | 146 | 0.42 | -32 | | -\$914,041 | 9.4 |
| Total | | | 253 | 37775 | 3870 | | 138 | | 1246 | | -326 | \$79,499,238 | -\$10,624,190 | 106.1 |

popn=population; Mort rate=mortality rate19 (per 100,000); Num FS=number of flexible sigmoidoscopies; Mort HR=mortality hazard rate; Dths prev=CRC deaths prevented; Inc rate CRC=incidence rate of CRC (per 100,000); Cases prev=cases of CRC prevented; Disc savings of treatment = discounted costs of treatment saved; Disc dths prev=discounted prevented deaths from CRC

The estimated cost of FS40 screening and subsequent surveillance colonoscopies was NZ$79,499,238. The long-term reduction in CRC incidence indicated that 326 members of the cohort would be prevented from developing CRC, resulting in savings of NZ$10,624,190 in the cost of cancer treatment over 20 years.

### FS45 screening

For screening by FS 45 in 2027 (Table 4), 200 (54%) of the 367 deaths expected from CRC were preventable. Each death prevented required 239 members of the cohort to be screened. Participation was assumed to be the same as for FS40.

**Table 4.** Twenty-year effect of screening by once-only flexible sigmoidoscopy at 45 years of age: participation 52.5% of which 5% recommended for colonoscopy of whom 25% require 3-yearly surveillance (costs given in NZ dollars).

| Age | Annual popn | Mort rate | Exp dths | Num FS | Colono-<br>scopies | Mort HR | Dths prev | Inc rate CRC | CRC w/o FS | Inc Distal HR | Cases treated | Disc cost of screening | Disc savings of treatment | Disc dths prev |
| --- | --- | --- | --- | --- | --- | --- | --- | --- | --- | --- | --- | --- | --- | --- |
| 45 | 71446 | 8.6 | 6 | 37585 | 1785 | 1 | 0 | 41.6 | 30 | 1.50 | 7 | \$72,594,932 | \$301,518 | 0.0 |
| 46 | 70265 | 9.4 | 7 | | | 0.87 | 2 | 45.5 | 32 | 0.21 | -11 | | -\$440,678 | 1.8 |
| 47 | 69084 | 10.2 | 7 | | | 0.74 | 4 | 49.4 | 35 | 0.12 | -13 | | -\$537,277 | 3.8 |
| 48 | 67903 | 12.5 | 9 | | 424 | 0.74 | 5 | 60.1 | 43 | 0.18 | -14 | \$1,087,354 | -\$573,572 | 4.6 |
| 49 | 66722 | 14.9 | 11 | | | 0.74 | 6 | 65.7 | 47 | 0.25 | -14 | | -\$551,390 | 5.4 |
| 50 | 65542 | 17.2 | 12 | | | 0.74 | 7 | 73.9 | 52 | 0.26 | -15 | | -\$559,654 | 6.2 |
| 51 | 64361 | 19.5 | 14 | | 403 | 0.74 | 8 | 82.0 | 58 | 0.20 | -18 | \$915,063 | -\$655,234 | 6.9 |
| 52 | 63180 | 21.9 | 16 | | | 0.74 | 9 | 90.2 | 64 | 0.20 | -19 | | -\$711,679 | 7.5 |
| 53 | 63723 | 24.1 | 17 | | | 0.74 | 10 | 107.1 | 76 | 0.24 | -23 | | -\$817,690 | 8.2 |
| 54 | 64266 | 26.4 | 19 | | 383 | 0.74 | 10 | 124.0 | 87 | 0.25 | -26 | \$770,072 | -\$929,460 | 8.7 |
| 55 | 64810 | 28.7 | 20 | | | 0.74 | 11 | 140.9 | 99 | 0.26 | -30 | | -\$1,020,929 | 9.3 |
| 56 | 65353 | 30.9 | 22 | | | 0.74 | 12 | 157.8 | 111 | 0.27 | -33 | | -\$1,095,829 | 9.8 |
| 57 | 65896 | 33.2 | 23 | | 364 | 0.74 | 13 | 174.7 | 123 | 0.28 | -32 | \$648,054 | -\$1,049,074 | 10.3 |
| 58 | 65793 | 34.3 | 24 | | | 0.74 | 14 | 192.3 | 135 | 0.31 | -38 | | -\$1,231,095 | 10.4 |
| 59 | 65690 | 35.4 | 25 | | | 0.74 | 14 | 209.8 | 147 | 0.29 | -40 | | -\$1,262,194 | 10.5 |
| 60 | 65586 | 36.5 | 26 | | 345 | 0.74 | 14 | 227.4 | 159 | 0.35 | -41 | \$545,370 | -\$1,286,750 | 10.6 |
| 61 | 65483 | 37.6 | 26 | | | 0.74 | 15 | 244.9 | 170 | 0.40 | -42 | | -\$1,280,763 | 10.7 |
| 62 | 65380 | 38.7 | 27 | | | 0.74 | 15 | 262.4 | 182 | 0.42 | -48 | | -\$1,440,162 | 10.7 |
| 63 | 65138 | 39.8 | 28 | | 328 | 0.74 | 16 | 279.9 | 194 | 0.38 | -51 | \$458,956 | -\$1,491,716 | 10.8 |
| 64 | 64897 | 40.9 | 28 | | | 0.74 | 16 | 297.4 | 206 | 0.42 | -45 | | -\$1,290,172 | 10.8 |
| Total | | | 367 | 37585 | 4032 | | 200 | | 2049 | | -546 | \$77,019,801 | -\$15,342,803 | 157.0 |

popn = population; Mort rate=mortality rate (per 100,000); Num FS=number of flexible sigmoidoscopies; Mort HR=mortality hazard rate; Dths prev=CRC deaths prevented; Inc rate CRC=incidence rate of CRC (per 100,000); Cases prev=cases of CRC prevented; Disc savings of treatment=discounted costs of treatment saved; Disc dths prev=discounted prevented deaths from CRC

**Table 5.** Twenty-year effect of screening by FIT 2-yearly (cut-off 75ng Hb/ml) from 45 of age: participation of 57.4% of which 7% initially recommended for subsequent colonoscopy of whom 25% require 3-yearly surveillance (costs given in NZ dollars).

| Age | Annual popn | Mort rate | Exp dths | Num FIT | Colono-scopies | Mort ratio | Dths prev | Inc rate CRC | CRC w/o FIT | IDR | Cases prev | Disc cost of screening | Disc savings of treatment | Disc dths prev |
| --- | --- | --- | --- | --- | --- | --- | --- | --- | --- | --- | --- | --- | --- | --- |
| 45 | 71446 | 8.6 | 6 | 20577 | 1368 | 1.00 | 0 | 41.6 | 30 | 2.23 | -21 | \$6,680,602 | \$267,051 | 0.0 |
| 46 | 70265 | 9.4 | 7 | 20577 | 1368 | 0.95 | 0 | 45.5 | 32 | 0.56 | 8 | \$6,546,990 | \$261,710 | 0.2 |
| 47 | 69084 | 10.2 | 7 | 19154 | 767 | 0.90 | 0 | 49.4 | 35 | 1.25 | -5 | \$4,562,607 | -\$31,164 | 0.4 |
| 48 | 67903 | 12.5 | 9 | 19154 | 767 | 0.85 | 1 | 60.1 | 43 | 0.73 | 7 | \$4,471,355 | -\$30,541 | 0.7 |
| 49 | 66722 | 14.9 | 11 | 18675 | 520 | 0.80 | 1 | 65.7 | 47 | 1.15 | -4 | \$3,665,193 | -\$266,281 | 1.1 |
| 50 | 65542 | 17.2 | 12 | 18675 | 520 | 0.75 | 2 | 73.9 | 52 | 0.40 | 18 | \$3,591,889 | -\$260,955 | 1.6 |
| 51 | 64361 | 19.5 | 14 | 18193 | 849 | 0.75 | 2 | 82.0 | 58 | 1.21 | -7 | \$4,306,630 | -\$247,465 | 1.7 |
| 52 | 63180 | 21.9 | 16 | 18193 | 849 | 0.75 | 2 | 90.2 | 64 | 0.44 | 20 | \$4,220,497 | -\$242,515 | 1.9 |
| 53 | 63723 | 24.1 | 17 | 17715 | 942 | 0.75 | 2 | 107.1 | 76 | 0.88 | 5 | \$4,311,177 | -\$88,917 | 2.1 |
| 54 | 64266 | 26.4 | 19 | 17715 | 942 | 0.75 | 3 | 124.0 | 87 | 1.00 | 0 | \$4,224,954 | -\$87,139 | 2.2 |
| 55 | 64810 | 28.7 | 20 | 17240 | 690 | 0.75 | 3 | 140.9 | 99 | 0.90 | 6 | \$3,493,548 | -\$204,899 | 2.3 |
| 56 | 65353 | 30.9 | 22 | 17240 | 690 | 0.75 | 3 | 157.8 | 111 | 0.90 | 6 | \$3,423,677 | -\$200,801 | 2.5 |
| 57 | 65896 | 33.2 | 23 | 16763 | 1014 | 0.75 | 3 | 174.7 | 123 | 0.90 | 7 | \$4,040,864 | -\$240,450 | 2.6 |
| 58 | 65793 | 34.3 | 24 | 16763 | 1014 | 0.75 | 3 | 192.3 | 135 | 0.90 | 8 | \$3,960,047 | -\$235,641 | 2.6 |
| 59 | 65690 | 35.4 | 25 | 16289 | 1101 | 0.75 | 4 | 209.8 | 147 | 0.90 | 8 | \$4,024,922 | -\$273,676 | 2.6 |
| 60 | 65586 | 36.5 | 26 | 16289 | 1101 | 0.75 | 4 | 227.4 | 159 | 0.90 | 9 | \$3,944,424 | -\$268,203 | 2.7 |
| 61 | 65483 | 37.6 | 26 | 15817 | 844 | 0.75 | 4 | 244.9 | 170 | 0.90 | 10 | \$3,281,894 | -\$303,370 | 2.7 |
| 62 | 65380 | 38.7 | 27 | 15817 | 844 | 0.75 | 4 | 262.4 | 182 | 0.90 | 10 | \$3,216,256 | -\$297,303 | 2.7 |
| 63 | 65138 | 39.8 | 28 | 15341 | 1163 | 0.75 | 4 | 279.9 | 194 | 0.90 | 11 | \$3,748,777 | -\$329,760 | 2.7 |
| 64 | 64897 | 40.9 | 28 | 15341 | 1163 | 0.75 | 4 | 297.4 | 206 | 0.90 | 12 | \$3,673,802 | -\$323,165 | 2.7 |
| Total | | | 367 | 351530 | 18514 | | 49 | | 2049 | | 107 | \$83,390,106 | -\$3,006,648 | 38.1 |

popn= population; Mort rate=mortality rate (per 100,000); Num FIT=number of faecal immunochemical tests; Mort ratio=mortality ratio; Dths prev=CRC deaths prevented; Inc rate CRC=incidence rate of CRC (per 100,000); CRC w/o FIT=CRC cases without FIT screening; IDR=Incidence
detection ratio; Cases prev=cases of CRC prevented; Disc cost of screening=discounted cost of screening; Disc savings of treatment= discounted costs of treatment saved; Disc dths prev=discounted prevented deaths from CRC

The long-term reduction in CRC incidence by FS45 resulted in savings of NZ$15,342,803 in the cost of cancer treatment.

### FIT75 screening

Biennial FIT75 screening from age 45 years prevented 49 deaths from CRC (13.3%) and 107 people developing CRC over 20 years. We estimated that 20.4% of the individuals in the FIT75 cohort would require screening by colonoscopy at some time over 20 years. Each premature death prevented required 1080 members of the cohort to be screened. The cost of screening was NZ$83,390,106 with relatively modest savings in treatment of NZ$2,104,653.

### FIT200 screening

For biennial FIT200 screening from age 45 (Table 6), 17.5% would have a colonoscopy. Biennial FIT200 prevented 41 deaths from CRC (13.3%) and 75 people from developing CRC. Each death prevented required 1274 members of the cohort to be screened.

**Table 6.** Twenty-year effect of screening by FIT 2-yearly (cut-off 200ng Hb/ml) from 45years of age: participation of 57.4% of which 5.1% initially recommended for subsequent colonoscopy of whom 25% require 3-yearly surveillance (costs given in NZ dollars).

| Age | Annual popn | Mort rate | Exp dths | Num FIT | Colono-scopies | Mort ratio | Dths prev | Inc rate CRC | CRC w/o FIT | IDR | Cases prev | Disc cost of screening | Disc savings of treatment | Disc dths prev |
| --- | --- | --- | --- | --- | --- | --- | --- | --- | --- | --- | --- | --- | --- | --- |
| 45 | 71446 | 8.6 | 6 | 20577 | 1982 | 1.00 | 0 | 41.6 | 30 | 1.86 | -15 | \$5,588,392 | \$186,936 | 0.0 |
| 46 | 70265 | 9.4 | 7 | 20577 | 0 | 0.96 | 0 | 45.5 | 32 | 0.69 | 6 | \$5,476,624 | \$183,197 | 0.2 |
| 47 | 69084 | 10.2 | 7 | 19531 | 627 | 0.92 | 0 | 49.4 | 35 | 1.18 | -4 | \$3,350,723 | -\$21,815 | 0.3 |
| 48 | 67903 | 12.5 | 9 | 19531 | 471 | 0.87 | 1 | 60.1 | 43 | 0.81 | 5 | \$3,283,708 | -\$21,379 | 0.6 |
| 49 | 66722 | 14.9 | 11 | 19164 | 615 | 0.83 | 1 | 65.7 | 47 | 1.11 | -3 | \$3,157,430 | -\$186,397 | 0.9 |
| 50 | 65542 | 17.2 | 12 | 19164 | 149 | 0.79 | 1 | 73.9 | 52 | 0.58 | 12 | \$3,094,282 | -\$182,669 | 1.3 |
| 51 | 64361 | 19.5 | 14 | 18787 | 1074 | 0.79 | 2 | 82.0 | 58 | 1.15 | -5 | \$2,972,872 | -\$173,225 | 1.5 |
| 52 | 63180 | 21.9 | 16 | 18787 | 146 | 0.79 | 2 | 90.2 | 64 | 0.61 | 14 | \$2,913,414 | -\$169,761 | 1.6 |
| 53 | 63723 | 24.1 | 17 | 18412 | 740 | 0.79 | 2 | 107.1 | 76 | 0.92 | 3 | \$2,798,023 | -\$62,242 | 1.8 |
| 54 | 64266 | 26.4 | 19 | 18412 | 614 | 0.79 | 2 | 124.0 | 87 | 1.00 | 0 | \$2,742,062 | -\$60,997 | 1.9 |
| 55 | 64810 | 28.7 | 20 | 17738 | 716 | 0.79 | 2 | 140.9 | 99 | 0.93 | 4 | \$2,588,857 | -\$143,429 | 2.0 |
| 56 | 65353 | 30.9 | 22 | 17738 | 289 | 0.79 | 3 | 157.8 | 111 | 0.93 | 4 | \$2,537,080 | -\$140,561 | 2.1 |
| 57 | 65896 | 33.2 | 23 | 17654 | 1181 | 0.79 | 3 | 174.7 | 123 | 0.93 | 5 | \$2,474,643 | -\$168,315 | 2.2 |
| 58 | 65793 | 34.3 | 24 | 17654 | 281 | 0.79 | 3 | 192.3 | 135 | 0.93 | 5 | \$2,425,151 | -\$164,949 | 2.2 |
| 59 | 65690 | 35.4 | 25 | 17268 | 844 | 0.79 | 3 | 209.8 | 147 | 0.93 | 6 | \$2,324,692 | -\$191,573 | 2.2 |
| 60 | 65586 | 36.5 | 26 | 17268 | 749 | 0.79 | 3 | 227.4 | 159 | 0.93 | 6 | \$2,278,198 | -\$187,742 | 2.3 |
| 61 | 65483 | 37.6 | 26 | 16880 | 823 | 0.79 | 3 | 244.9 | 170 | 0.93 | 7 | \$2,182,405 | -\$212,359 | 2.3 |
| 62 | 65380 | 38.7 | 27 | 16880 | 421 | 0.79 | 3 | 262.4 | 182 | 0.93 | 7 | \$2,138,757 | -\$208,112 | 2.3 |
| 63 | 65138 | 39.8 | 28 | 16483 | 1278 | 0.79 | 3 | 279.9 | 194 | 0.93 | 8 | \$2,046,747 | -\$230,832 | 2.3 |
| 64 | 64897 | 40.9 | 28 | 16483 | 410 | 0.79 | 3 | 297.4 | 206 | 0.93 | 8 | \$2,005,813 | -\$226,215 | 2.3 |
| Total | | | 367 | 364987 | 12420 | | 41 | | 2049 | | 75 | \$58,379,872 | -\$2,104,653 | 32.3 |

popn= population; Mort rate=mortality rate (per 100,000); Num FIT=number of faecal immunochemical tests; Mort HR=mortality hazard rate; Dths prev=CRC deaths prevented; Inc rate CRC=incidence rate of CRC (per 100,000); CRC w/o FIT=CRC cases without FIT screening; IDR=Incidence detection ratio; Cases prev=cases of CRC prevented; Disc cost of screening=discounted cost of screening; Disc savings of treatment= discounted costs of treatment saved; Disc dths prev=discounted prevented deaths from CRC

### Relative impact and cost of different screening approaches

The discounted health-service savings from treatment of CRC avoided because of reduced annual incidence were subtracted from the cost of screening and subsequent colonoscopies and divided by the discounted number of CRC deaths prevented to provide estimates the screening and treatment cost-effectiveness ratio of the effect of screening on the emerging CRC mortality epidemic (Table 7). The continuing expense of screening, treatment, and subsequent colonoscopies suggested that FIT200 or FIT75 screening would cost more and prevent many fewer deaths than one-off FS screening. One-off FS45 screening prevented most deaths from CRC and was the most cost-effective of the screening scenarios assessed and produced the largest government taxation gained and survivor’s benefit avoided by the prevention of mortality in those aged 45-64 years.

**Table 7.** Discounted health and other government costs of screening and treatment over 20 years (costs given in NZ dollars).

| Screening strategy | Discounted screening & treatment cost | Discounted CRC deaths prevented | Cost per death prevented | Discounted tax gained | Survivor's benefit saved |
| --- | --- | --- | --- | --- | --- |
| One-off FS at age 40 | \$68,875,048 | 106.1 | \$649,161 | \$9,869,836 | \$5,384,147 |
| One-off FS at age 45 | \$59,096,001 | 157.0 | \$376,408 | \$9,040,823 | \$8,081,603 |
| FIT75 from age 45 | \$83,390,106 | 38.1 | \$2,188,717 | \$2,009,473 | \$1,956,099 |
| FIT200 from age 45 | \$53,892,780 | 32.3 | \$1,668,507 | \$1,705,007 | \$1,659,721 |

## Discussion

Strongly influenced by changes in risk among different birth cohorts,^1,2^ CRC is rising among generations aged <50 years in several countries.^2,3^ There is a need to improve early diagnosis of YOCRC as well as screening among those <50 years; the NBSP does not currently cover this at-risk group. In ANZ, a two-stage system is used whereby the candidates for screening colonoscopy are identified among those 60-75 years using FIT200 to detect haemoglobin in the stool. Unfortunately, ANZ has a poor history of managing relevant services to cope with the increased need imposed as a result of cancer screening;^23^ delays of up to 10 months between the onset of symptoms and diagnosis have been reported.^24^ Although colonoscopy is recommended for the investigation of patients with rectal bleeding or persistent anaemia, FS as the initial approach can greatly reduce the demand for colonoscopy when services are unable to keep up with need.^6–8^

A partial response to the emerging epidemic would involve lowering the NBSP starting age for screening. Improving diagnostic responsiveness and lowering the age at commencing screening both involve greater endoscopy resources. A rational response would be the expansion of FS services.

One-off FS at age 40 or 45 would prevent more than 50% of deaths from CRC over 20 years, compared with approximately 13% for FIT75 and FIT200. The cost savings for FS 40 and FS45 were estimated at approximately NZ$10 million, and NZ$15 million, respectively. FS 45 was shown to prevent most deaths from CRC and to be the most cost-effective screening strategy. ^14,15^

When screening largely enables effective treatment of pre-invasive disease, it can have a pronounced long-term effect on incidence, as observed in FS screening.^25^ When prevalent cases present at the procedure are excluded, one-off FS has been shown to produce a 66% (95%CI: 61%-70%) reduction in distal CRC and a 41% (95%CI: 35%-45%) reduction in overall CRC incidence,^26^ Consistent with these findings, a meta-analysis of various screening modalities has found that it is likely that only FS increases overall life expectancy.^27^ Simulation models comparing screening programmes have underestimated^27^ the per-protocol reduction in mortality and incidence achievable and have greatly underestimated the cost-effectiveness of one-off FS screening over the long term.^17,27^

Because of serious under-resourcing of endoscopy services in ANZ and growing concern with the high incidence of YOCRC, at the request of the local District Health Board one of us (PB) investigated adults <50 years with monosymptomatic fresh rectal bleeding; it was undertaken by initial FS. Between 2017 and end of 2023, 1389 patients had FS and 1.1% had CRCs, 27.4% colorectal polyps and 4.9% high risk lesions.^29^ Hence, timely FS is an appropriate clinical response for people with symptoms of YOCRC when the need for colonoscopy exceeds capacity.

The analysis presented here confirms that one-off FS screening at 45 years of age would provide the most immediate, long-lasting, and cost-effective response to the emerging epidemic of YOCRC.

The higher proportion of participants requiring colonoscopy in FIT scenarios results in more hospitalisations than for one-off FS screening due to the greater number of colonoscopies performed over 20 years.^13^

In several countries, FS is performed by appropriately trained nurses, allowing timely endoscopic investigation of patients with symptoms of YOCRC. A 7-month FS <u>training</u> <u>course</u> for nurses and other health professionals was established in the UK more than 10 years ago by the Joint Advisory Group on Gastrointestinal Endoscopy (JAG) with an optional path for those who wish to complete full endoscopy training. Long-term benefit would accrue to ANZ from its adoption and would upskill and extend the ANZ endoscopy workforce so that the additional screening needed in the face of the emerging epidemic of YOCRC and the management of symptomatic patients with gastroenterological symptoms does not compete with the completion-colonoscopy needs of the NBSP.^30^

## Data Availability

All data produced in the present work are contained in the manuscript

https://tewhatuora.shinyapps.io/mortality-web-tool/

https://www.stats.govt.nz/information-releases/national-population-projections-2022base2073/

https://www.healthpoint.co.nz/private/gastroenterology/the-rutherford-clinic/

https://www.alleviaradiology.co.nz/our-services/screening-programmes/bowel-screening

https://www.ird.govt.nz/about-us/tax-statistics/revenue-refunds/income-distribution

https://www.ird.govt.nz/about-us/tax-statistics/revenue-refunds/income-distribution

